# Peripheral innate immunophenotype in neurodegenerative disease: blood-based profiles and links to survival

**DOI:** 10.1101/2024.01.16.24301348

**Authors:** Alexandra Strauss, Peter Swann, Stacey Kigar, Rafailia Christou, Natalia Savinykh Yarkoni, Alexander Murley, Leonidas Chouliaras, George Savulich, Richard Bevan-Jones, Ajenthan Surendranthan, John O’Brien, James Rowe, Maura Malpetti

## Abstract

The innate immune system plays an integral role in the progression of many neurodegenerative diseases. In addition to central innate immune cells (e.g. cerebral microglia), peripheral innate immune cells (e.g. blood monocytes, natural killer cells, and dendritic cells) may also differ in these conditions. However, the characterization of peripheral innate immune cell types across different neurodegenerative diseases remains incomplete. This study aimed to characterize peripheral innate immune profiles using flow cytometry for immunophenotyping of peripheral blood mononuclear cells, in n=148 people with Alzheimer’s disease (AD), Frontotemporal Dementia (FTD), Corticobasal syndrome (CBS), Progressive Supranuclear Palsy (PSP), Lewy Body Disease (LBD) as compared to n=37 healthy controls. To compare groups, we used Principal Component Analysis and multivariate Dissimilarity analysis across 19 innate immune cell types. We identified pro-inflammatory profiles that significantly differ between patients with all-cause dementia and healthy controls, with some significant differences between groups. Regression analysis confirmed that time to death following the blood test correlated with the individuals’ immune profile weighting, positively to TREM2+ and nonclassical monocytes and negatively to classical monocytes. Taken together, these results describe transdiagnostic peripheral immune profiles and highlight the link between prognosis and the monocyte cellular subdivision and function (as measured by surface protein expression). The results suggest that blood-derived innate immune profiles can inform sub-populations of cells relevant for specific neurodegenerative diseases that are significantly linked to accelerated disease progression and worse survival outcomes across diagnoses. Blood-based innate immune profiles may contribute to enhanced precision medicine approaches dementia, helping to identify and monitor therapeutic targets and stratify patients for candidate immunotherapies.

## Introduction

Neurodegenerative diseases are an increasingly common aspect of the ageing population globally, with a pressing need for new preventive and disease-modifying therapies. Despite the clinico-pathological heterogeneity of neurodegenerative diseases, and diversity of amyloid, tau and alpha-synuclein proteinopathies, they share a dysregulation of the innate immune system features in all dementias (1). Centrally, activation of cerebral microglia has been reported for Alzheimer’s disease (AD) and its prodromal mild cognitive impairment (MCI), Lewy Body Disease (LBD) including Parkinson’s disease dementia (PDD) and Dementia with Lewy Bodies (DLB), and the syndromes associated with frontotemporal lobar degeneration (FTLD). The latter include the behavioral variant of frontotemporal dementia (bvFTD), non-fluent (nfPPA) and semantic (svPPA) variants of primary progressive aphasia, corticobasal syndrome (CBS), and progressive supranuclear palsy (PSP). Moreover, genome-wide association studies link mutations in genes coding for proteins related to the immune system to the development of multiple neurodegenerative conditions (2–7) Mutations in the gene for triggering receptor expressed on myeloid cells 2 (TREM2, encoding a receptor on monocytes and microglia) may cause FTLD-syndromes and AD, with an effect size comparable to the immune-related APOE4 variant (2,8). Such genomic associations are complemented by evidence for abnormalities in central and peripheral innate immune systems in neurodegenerative diseases (9).

Most evidence describing abnormal innate immunity in dementia to date concerns cerebral cells. For example, microglial activation in the central nervous system is implicated in many forms of dementia (10–16). For example, human post mortem studies report activated microglia in association with the severity of amyloid and tau pathology in AD, FTLD-syndromes and LBD (11,12,17,18). However, post mortem studies are not well suited to characterize microglial changes in early disease stages. Instead, overexpression of the translocator protein 18 kDa (TSPO), overexpressed in activated microglia, can be detected by positron emission tomography (PET). Microglial activation presents early in people living with many types of dementia and predicts the rate of cognitive decline (13–16,19). While TSPO PET is a powerful, informative tool and relates directly to cerebral innate immune activation it is expensive and not readily scalable. Alternatively, peripheral blood-based markers of immune dysregulation would potentially be scalable and repeatable over time.

Cells of the innate immune system—including monocytes, dendritic cells, and natural killer (NK) cells – rapidly and non-specifically initiate an immune response upon detection of pathogens or cellular damage (20). While this process is beneficial in the short-term to eliminate harmful stimuli, chronic or dysregulated activation can lead to disease. The peripheral immune system may interact with the central nervous system indirectly via chemokine and cytokine signaling or directly via infiltration into and meningeal surveillance of the parenchyma (21–27). Elevated pro-inflammatory cytokine concentrations are reported in post mortem brain tissue and cerebrospinal fluid (CSF) in people with AD, LBD, and FTLD-syndromes (28–31); while peripheral infections can exacerbate the neuroinflammatory environment and accelerate cognitive decline (26,32). Taken together, evidence implicates primary effector cells of the peripheral innate immune system in the pathogenesis of neurodegenerative diseases.

Monocytes, natural killer (NK) cells, and dendritic cells have each been implicated in the pathogenesis of dementias. For example, TREM2 is expressed on the surface of peripheral monocytes (33) and may be a marker of monocyte recruitment in AD (27). Elevated concentration of CSF soluble TREM2 (sTREM2), cleaved from the membranes of microglia or monocytes, is linked to disease progress cognitive decline in AD (34,34) and FTD (11). Beyond TREM2, CSF concentrations of chemokine motif ligand 2 (CCL2), mediating chemotaxis of monocytes, is also predictive of cognitive decline in AD (35). Similarly, abnormal activation patterns of NK cells occur with AD (36), Parkinson’s disease (closely related to DLB and PDD) (37). Finally, myeloid dendritic cells are reduced in people with AD and Parkinson’s disease (38–40). Overall, evidence involving monocytes, NK cells, and dendritic cells supports a more integrative disease processes than initially appreciated.

While the mechanisms of action of peripheral immune cells remain unclear, they are nonetheless more readily accessible to quantify, compared to central innate immune cells, via phlebotomy. The identification of innate cell types in the periphery of people with dementia may clarify links between innate immunity and neurodegenerative disease, and yield clinically relevant, blood-based biomarkers to support target identification and patient stratification for immune-targeting therapies. However, current immunophenotyping data is limited to few diagnoses of dementia and remains incomplete in cellular subtype characterization (38,39,41–43). To meet the needs for diagnostic, prognostic and trialist use, there is a pressing need for improved profiling of peripheral innate immunity in multiple neurodegenerative disorders.

This study therefore aims to characterize innate immune profiles in blood of people with diverse neurodegenerative dementias, including AD, FTLD-syndromes and LBD using Flow Cytometery-a well-established method of identifying cells via their unique expression of activation and lineage markers on the cell surface. We test the hypothesis that innate immune cell profiles are abnormal in people with any of these disorders, with at least partial commonality in the immune profile for the neurodegenerative disorders investigated here (i.e. a transdiagnostic abnormality). Given the multivariate nature of immune activation, we use data driven approaches to reduce dimensionality (complexity) and test for (dis)-similarity between the clinical diagnostic groups. We then highlight clinical relevance by the correlation or multivariate immune profiles with time to death.

## Materials and Methods

### Participants

Patients (n =148) were recruited from clinics for cognitive and movement disorders at the Cambridge University Hospitals NHS Trust as well as collaborating regional psychiatry and neurology services. We included participants with a clinical diagnosis of MCI/AD (44,45) (n=24), probable or possible PSP (n=54, predominantly Richardson syndrome phenotype), CBS (46)(n=22), bvFTD/PPA (47)(n=19), PDD (n=3), or DLB (n=27) (48,49). Of the CBS patients, nine underwent amyloid PET, which confirmed amyloid-positivity in three. The MCI/AD cohort comprised sixteen people with AD and five with MCI. Of the MCI patients, three out of five underwent amyloid PET and all were confirmed amyloid positive. The FTD cohort comprised thirteen people with behavioral variant FTD (bvFTD) and six people with PPA. The LBD cohord comprised twenty-seven people with DLB and three with PDD. We included thirty-seven healthy controls with MMSE > 26/30, absence of memory symptoms, no signs of dementia, or any other significant medical illnesses. For participants who did not have the mental capacity to consent, we followed the personal consultee process as set out by UK law. For participants with mental capacity, all gave written informed consent according to the Declaration of Helsinki. The study protocol was approved by the NIHR National Research Ethic Service Committee and East of England (Cambridge Central).

Participants underwent baseline clinical and neuropsychological assessment, including the revised Addenbrooke’s Cognitive Examination (ACE-R, 0-100 points) and mini-mental state examination (MMSE). The ACE-R is subdivided into five domains: Attention and Orientation (18/100), Memory (26/100), Fluency (14/100), Language (26/100), and Visuospatial abilities (16/100). Survival data were collected for all up to and including the 23^rd^ of October 2023 (the census date).

### Blood collection and flow cytometry analyses

At baseline, 18 ml of blood was drawn in EDTA and analyzed at the NIHR Cambridge Cell Immunophenotyping Hub. Wherever possible, samples were processed within 2h (over 98% of samples). Blood was layered onto sterile Ficoll (Cytiva, Cat#: 17144003) for peripheral blood mononuclear cell (PBMC) isolation by a technician blind to group status. Two aliquots containing ∼1 x10^6 PBMCs were stained using the antibody cocktail shown in Supplementary Table S2, using either TREM2 or a matched isotype control. At the end of staining, cells were washed, and the data were acquired on live (i.e., non-fixed) cells with a BD LSR Fortessa instrument.

Cell classifications were determined via manual gating in FlowJo (BD) by individuals blind to participant group according to standards recommended for the Human Immunology Project (50). Briefly, monocytes were identified as HLA-DR+/CD14+; populations were further stratified based on CD16 and CD14 positivity where CD16−/CD14hi monocytes were labelled as classical, CD16+/CD14+ monocytes labelled as intermediate, and CD16+/CD14lo monocytes labelled as nonclassical. Each monocyte subpopulation was then gated to discern CCR5+, CCR2+, and TREM2+ staining. Dendritic cells (DCs) were identified as HLA-DR+/CD14- and further stratified based on CD123 and CD11c staining: CD123+/CD11c+ cells were double positive (DC+/+), CD123+/CD11c− were plasmacytoid DCs, CD123−/CD11c+ were myeloid DCs, and CD123−/CD11c− were double negative (DC−/−). NK cells were identified as CD56+ and further subdivided into two groups as either CD16+ (which were CD56lo) or CD16− (which were CD56hi). Interrater validation studies showed excellent concordance between different evaluators (Supplementary Table S3.).

### Statistical Analysis

Analyses were performed using R (Version 2023.03.0+386). Non-parametric tests were used for all pairwise comparisons given non-normal data distributions. For inference, p-values were corrected for multiple comparisons using the Benjamini-Hochberg procedure to control the false discovery rate (FDR) and considered significant with threshold p-FDR < 0.05. For transparency and explorative analyses, uncorrected p-values are also reported. Statistical analysis was carried out in three steps.

First, to understand similarity of innate immune cell profiles between diagnoses and controls, absolute cell numbers from FlowJo were normalized to standardize across minimum and maximum absolute cell count ranges. These normalized cell counts were averaged within each diagnosis to yield an average, normalized 19-cell vector per diagnosis. To evaluate dissimilarity between diagnoses, Euclidean distance was calculated on the normalized data based on a 19-cell vector for all diagnoses followed by complete linkage hierarchical clustering.

Second, a Principal Component Analysis (PCA) was applied on the 19 innate immune cell classes. This reduced the dimensionality of our dataset to minimize multiple comparisons, identifying a limited number of components that best explain the data variance. The PCA was computed using cell counts calculated as a proportion of their parent population across all cell types (Supplementary Figure S1). Local Outlier Factor (LOF) analysis with a hyperparameter of k=10 identified 2 participants as outliers following PCA computation (LOF = 3, Supplementary Figure S5) (51). These outliers were excluded before re-computing the PCA. We retained 3 components based on eigenvalues >1, explained variance > 10 %, and application of the visual “elbow method” from the scree plot for further analyses (52,53) (Supplementary Figure S4). Individual participant loadings in each of the 3 selected components were included in group-comparison analyses using Mann Whitney U tests to compare median values between patients and controls, and Kruskal Wallace tests to compare across patient groups followed by Dunn’s post hoc analysis, where applicable.

Finally, we tested for associations between the individual participant loadings and clinical outcome. Individual component loadings were evaluated as a predictor of ACE-R total scores using a mixed linear regression followed by Spearman’s correlation analysis, controlling for age and sex. Similarly, we investigated each principal component loading in relationship to survival years using a linear model.

## Results

Participant summary clinical characteristics are in Table 1. There were 74 female and 109 male participants. Most groups, apart from CBS, had more males than females. As expected, control participants scored significantly higher on the ACE-R examination than each patient group (p < 0.001 for all comparisons); while people with AD had significantly lower ACE-R total scores than people with PSP (p = 0.002).

**Table 1.**
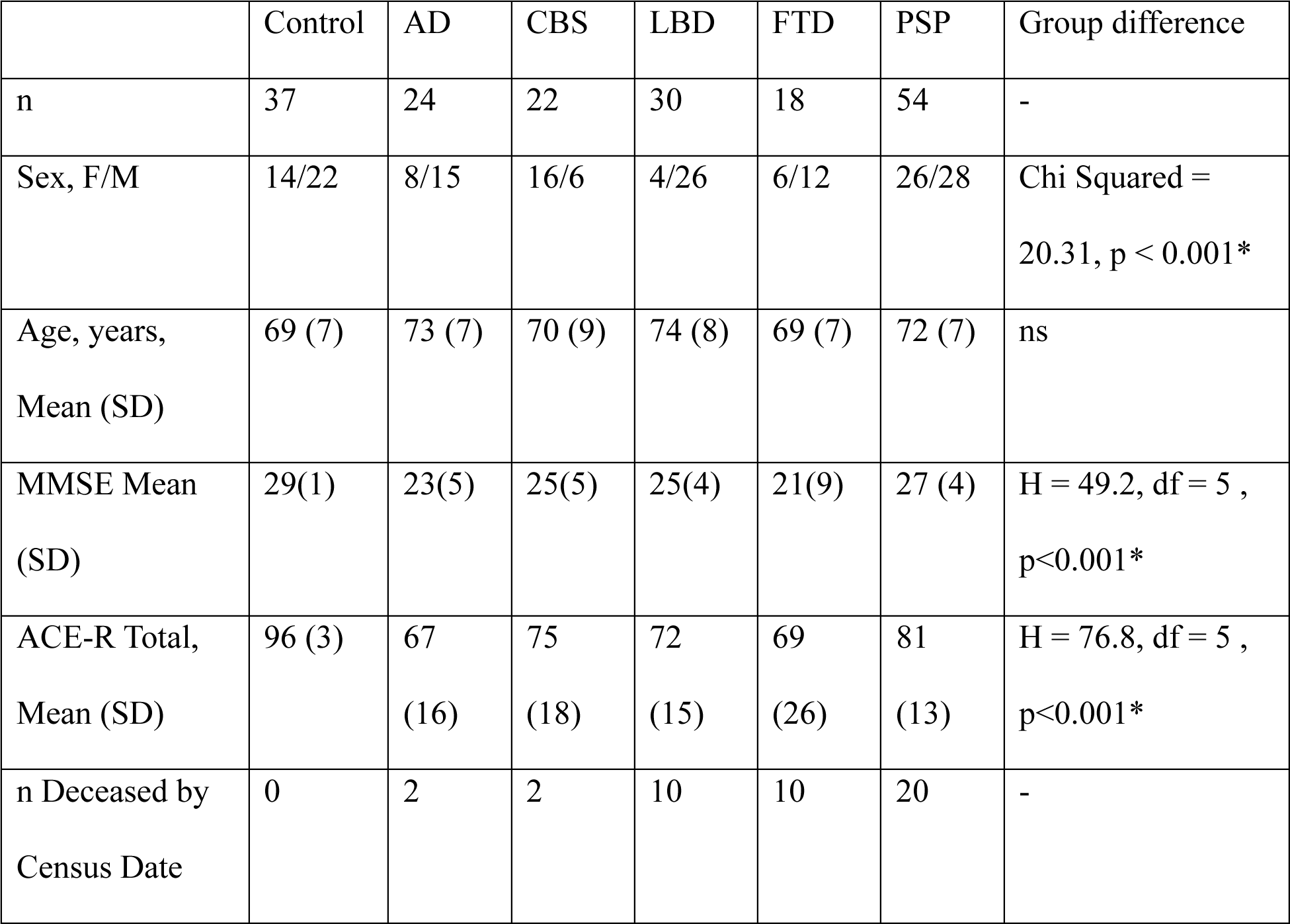
Demographic Information and cognitive test results for controls and patients in the study. AD = Alzheimer’s disease, CBS = Corticobasal syndrome, LBD = Lewy Body Disease, FTD = frontotemporal dementia, PSP = progressive supranuclear palsy. Differences in sex were evaluated using Chi Squared test. Differences in cognitive test scores were evaluated using Kruskal Wallace Tests followed by Dunn’s Post hoc analysis giving H. * indicates statistical significance between groups at p < 0.05.

### Dissimilarity Analysis

Figure 1 shows dissimilarity values of the 19-cell vector measured by Euclidean distance between groups (See Supplementary Figure S6 for individual cell pairwise comparisons). Hierarchical clustering represented in the dendrogram summarises the group-wise differences in the pattern (rather than magnitude) of immune profiles. Maximal distance was found between controls and all patient groups. The smallest distance (i.e. most similarity) was found between PSP vs CBS groups, while PSP/CBS and FTD groups were more similar to each other than to AD/LBD.

**Figure 1.**
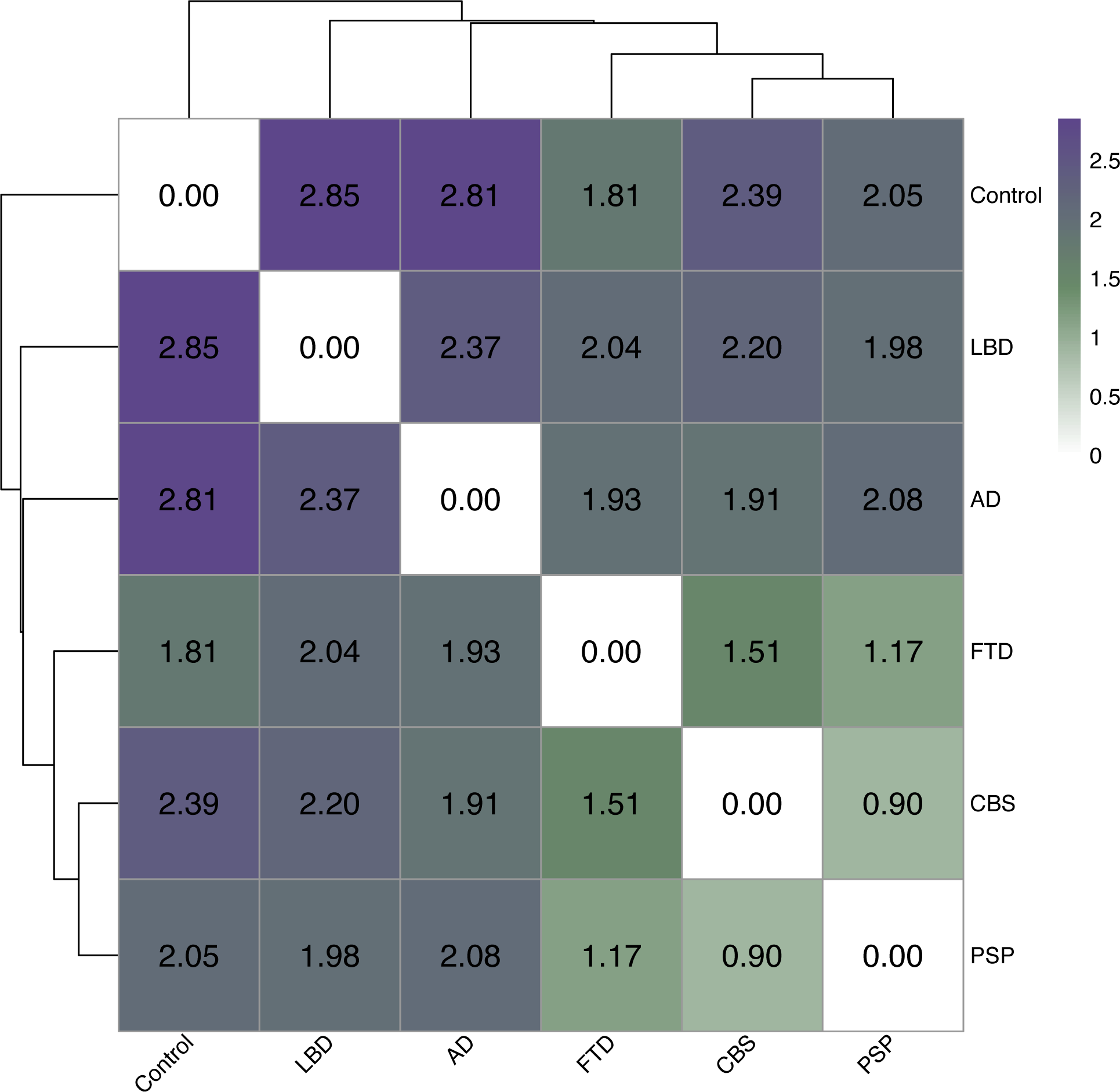
Euclidean Distance dissimilarity analysis and hierarchical clustering. Darker colors and larger values indicate greater dissimilarity between corresponding diagnostic groups, while lighter colors indicate relative similarity. In the dendrogram, note that all patients separate from controls initially, and that the family of disorders associated with frontotemporal lobar degeneration pathologies are relatively similar to each other, in contrast to Alzheimer’s disease and Dementia with Lewy bodies.

### Principal Component Analysis

From the PCA on the 19 cell populations, three components were selected for further analyses. Together, these components accounted for half of the total variance in the data (Supplementary Table S5). Shows the contribution of each cell type to the selected components. The first principal component (PC1) accounted for 18.2 % of the variance and was strongly positively weighted by TREM2+ monocytes and nonclassical monocytes (excluding CCR5+ nonclassical monocytes). PC1 was most negatively loaded onto classical monocytes, including CCR2+ classical monocytes, as well as NK cells high in CD16 expression (CD16+ NK cells) (Figure 2C). The second principal component (PC2) accounted for 16.6 %of the total variance. PC2 was strongly, negatively weighted by intermediate monocytes and CCR5+ monocytes, and strongly positively weighted by dendritic cells negative for CD11c and CD123 (Figure 3C). The third principal component (PC3) accounted for 12.8 % of the total variance and was most strongly positively weighted by CD16− NK Cells, and most strongly, negatively weighted in CD16+ NK cells and TREM2+ monocytes (Figure 4C). PC relationships to sex and age are found in Supplementary Table S6. Regression analysis revealed that there was no significant relationship between individual loadings in PC1, PC2, or PC3 and age (p >.05). There was no significant relationship between individual loadings in PC1 or PC2 and sex. Females had significantly lower median loading in PC3 as compared to males (p = .031, Females = −0.28, Males = 0.126).

**Figure 2.**
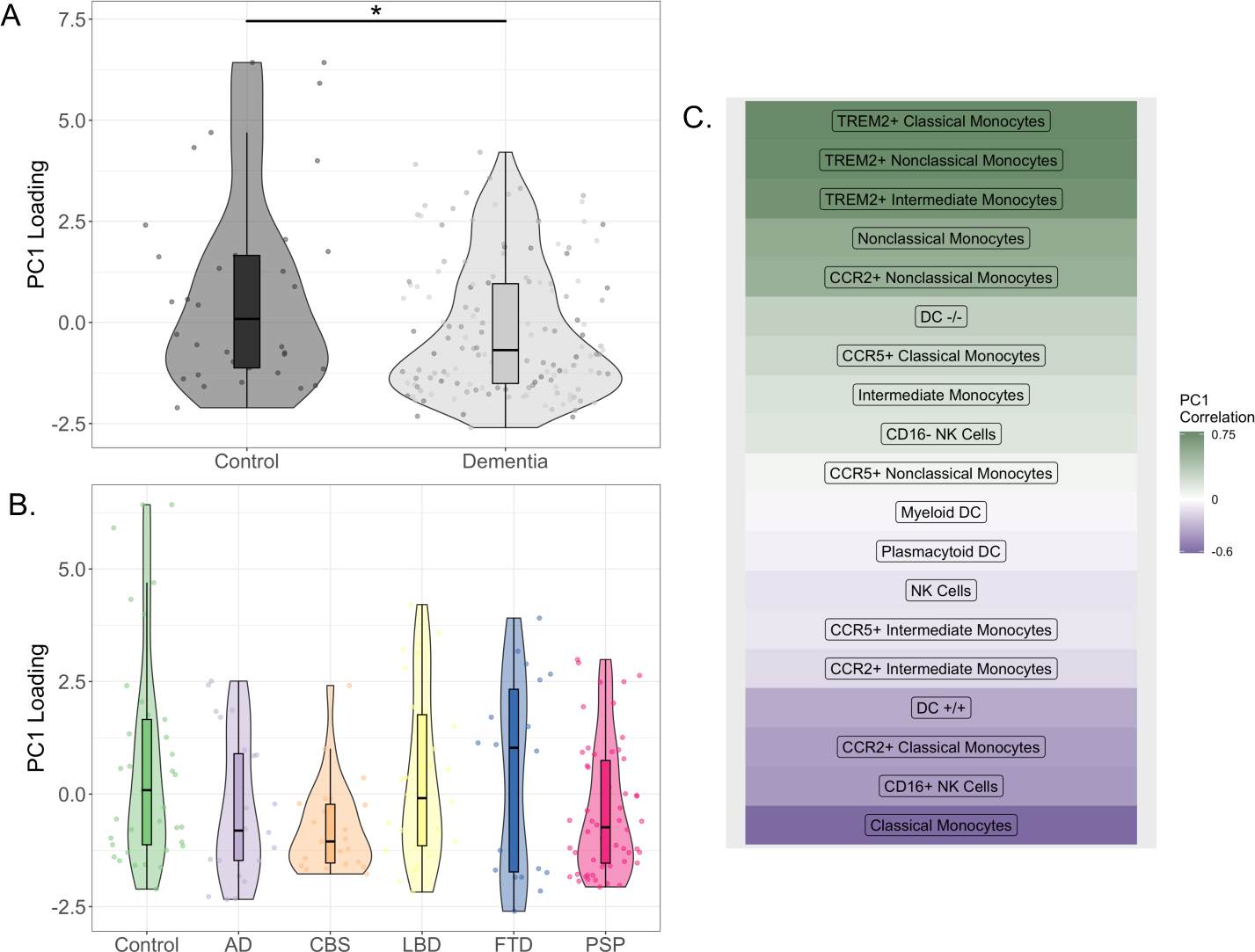
A) Median individual loading onto PC1 between controls and all-cause dementia patients, p = 0.02. B) Median individual loading onto PC1 for each diagnostic group. Note the absence of significant differences between patient groups. C) Cellular correlations to PC1 extracted following PCA. Darker colors indicate stronger positive and negative correlations to PC1. Mann Whitney U and Kruskal Wallace tests were used to compare medians. Results were considered significant at p < .05 indicated with *. A dashed line indicates a result of statistical significance prior to FDR correction that did not maintain significance following correction.

**Figure 3.**
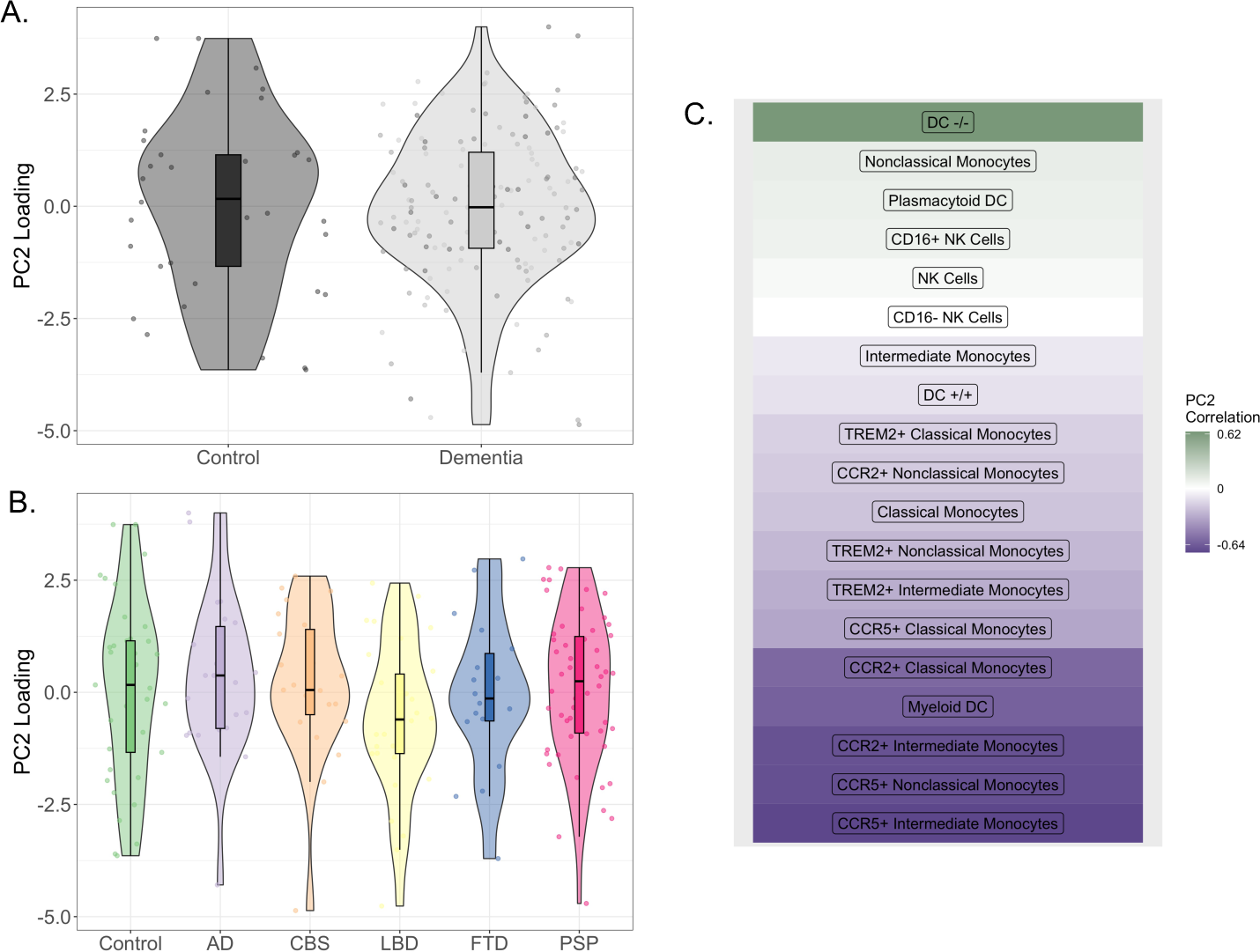
A) Median individual loading onto PC2 between controls and all-cause dementia patients. B) Median individual loading onto PC2 for each group. C) Cellular correlations to PC2 extracted following PCA. Darker colors indicate stronger positive and negative correlations to PC3. Mann Whitney U and Kruskal Wallace tests were used to compare medians. Results were considered significant at p <0.05.

**Figure 4.**
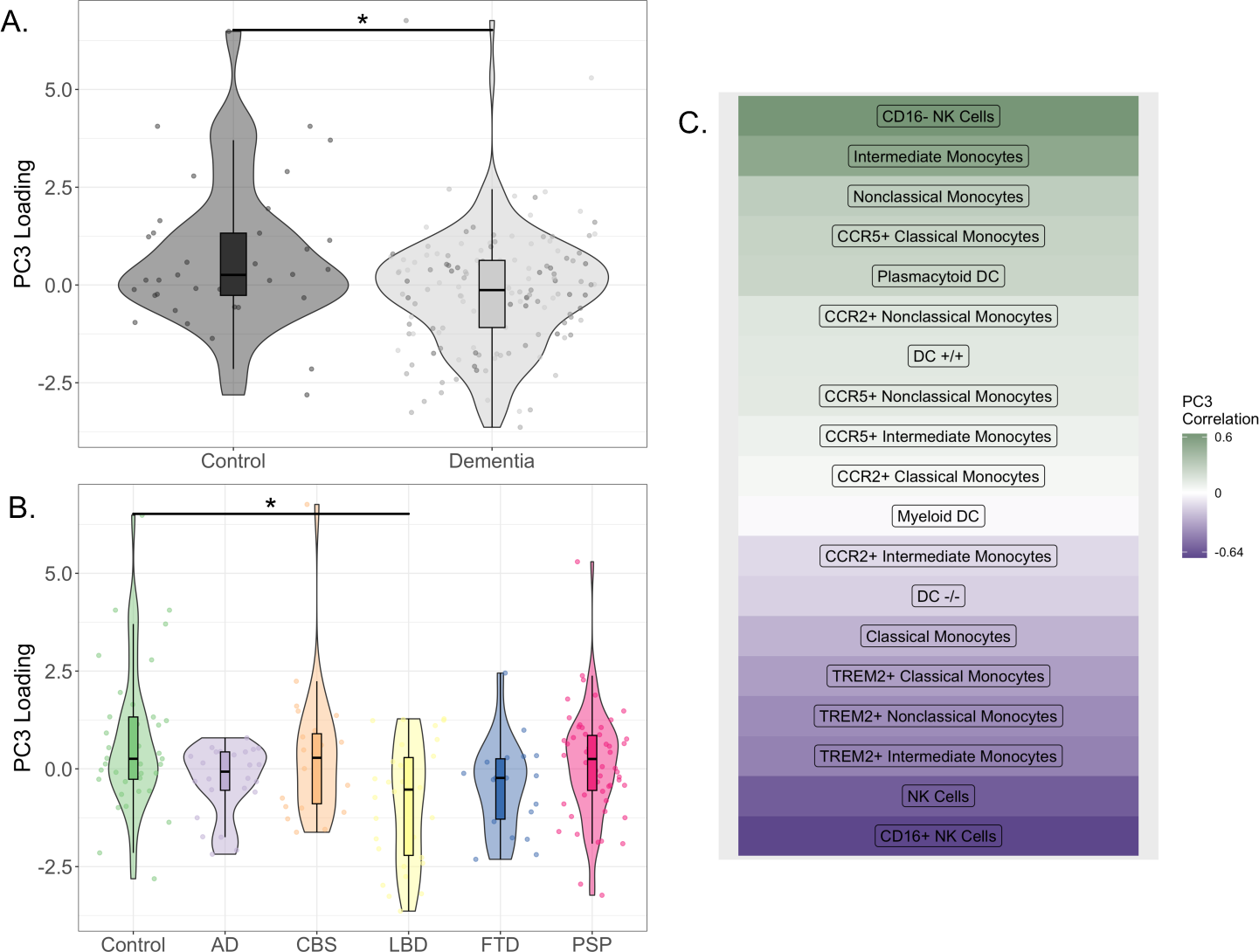
A) Median individual loading onto PC3 between controls and all-cause dementia patients, p = 0.015. B) Median individual loading onto PC3 for each group. Post hoc comparisons indicate LBD differs individually from controls, p = 0.01. C) Cellular correlations to PC3 extracted following PCA. Darker colors indicate stronger positive and negative correlations to PC3. Mann Whitney U and Kruskal Wallace tests were used to compare medians followed by Dunn’s Post hoc analysis with FDR correction for multiple comparison.

All-cause dementia patients showed significantly lower median loading values in PC1 (U = 3377, p= 0.034) as compared to controls (Figure 2A). Multi-group comparison did not identify a significant difference in PC1 loading between controls and each patient group (H= 9.88, p = 0.078) (Figure 2B). There was no statistically significant difference in individual PC2 loading between all-cause dementia and controls (U= 2844, p =0.767) or between controls and each patient group (H = 4.88, p = 0.481) (Figure 3A and B). All-cause dementia patients had significantly reduced PC3 loading as compared to controls (U= 2041, p=0.0147) (Figure 4A). Multi-group comparison identified a significant difference in PC3 loading across groups (H= 16, p= 0.007), and post hoc analysis revealed that patients with LBD showed significantly lower median loading values in PC3 than controls (p-FDR = 0.01) (Figure 4B). See supplementary Table S4 for statistics from post hoc analysis group comparisons.

### Clinical Outcomes

Individual patient loadings of PC1, PC2, and PC3 were each evaluated as a predictor of ACE-R scores including the interaction of diagnostic group as an independent variable. Regression analysis did not reveal significant predictive value of PC1, PC2, or PC3 for ACE-R total scores. There was no interaction of diagnostic group, age, or sex, with any principal component in terms of ACE-R score.

We used our data to evaluate the sample size required for traditional Cox proportional hazards regression. We completed a power calculation (power = 0.8, alpha = 0.05). To detect a hazard ratio of 1 +/− 0.2 as seen in previous studies using this regression in immune data in dementia (50), we would require 234 patients with confirmed death, or 835 total participants given our current distribution of deceased to living patients (28%). To carry out a primary survival analysis, we tested whether PC1, PC2 or PC3 were predictive of years of time to death in the subset of 42 patients who died prior to our census date (2 AD, 2 CBS, 10 LBD, 8 FTD, and 20 PSP). Higher individual loadings in PC1 predicted longer time from the research visit to death (*y*=0.4501*x*+2.0821, F = 10.098, p = 0.0028, *r*_2_=0.1816). This correlation remained after controlling for age, sex, and diagnosis (F = 3.689, p = 0.0066, *β_Age_*= −0.34, *β_Male_*=0.20). There was no association between PC2 (F = 0.019, p = 0.9) or PC3 (F = 2.734, p = 0.106) and years of survival following blood draw.

**Figure 5.**
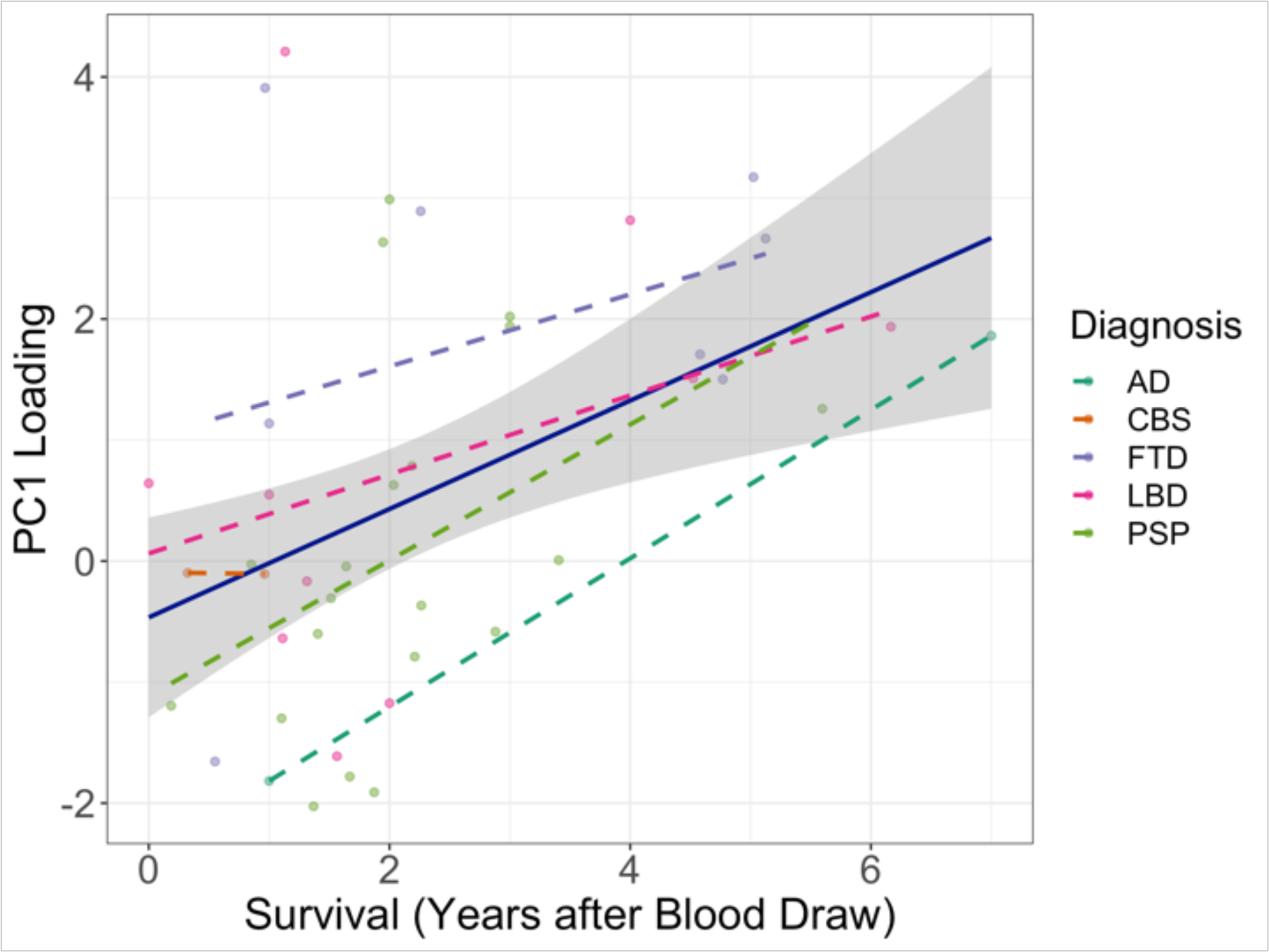
Years of survival following blood draw against individual PC1 loading across the subset of 42 patients who had died by the census date. A linear regression was used to establish how years of survival following blood draw was predicted by individual PC1 loading across all patient groups (*y* = 0.4501*x* + 2.0821, F = 10.098, p = 0.0028, *r*_2_ = 0.1816). Outcomes were considered significant at p < .05.

## Discussion

The main outcome of this study is confirmation that peripheral blood-based innate immunophenotypes are abnormal in people with AD, LBD, FTD, PSP, and CBS. The principal components of the 19-cell class immunophenotype were similarly abnormal in each type of neurodegenerative disease, and one component (or immune profile) predicted time to death. We identified transdiagnostic similarly in the *magnitude* of this abnormality in PC1, and diagnostic specificity in the magnitude and composition of this abnormality for LBD patients in PC3. Importantly, the multivariate *pattern* of cell-types was dissimilar between controls and all-cause dementia participants in this study. Taken together, these results indicate that even peripheral immune profiles have diagnostic specificity and identify a cellular profile represented by a redistribution of monocyte subtypes as being linked to survival across neurodegenerative dementias.

In the introduction, we proposed that peripheral innate immune cells are directly related to the pathogenesis of neurodegenerative diseases, based on genomics data and the physiological integration between central and peripheral immune compartments (2–7,38–43,54,55). Rather than a cell-by-cell account, we adopted a multivariate approach and identified three principal components of interest. The first (PC1) was weighted positively to TREM2+ and nonclassical monocytes and negatively to classical monocytes. The second (PC2) was weighted positively to CCR5+ monocytes and intermediate subtypes, and negatively to DC−/− cells. The third component (PC3) was positively weighted by CD16− NK cells and negatively weighted by CD16+ NK cells. These multivariate (multi-cell type) patterns help to contextualize the results of more selective prior studies.

For example, the finding of altered monocyte subpopulations in AD that varied with disease stage (41), albeit without natural killer cell or dendritic cell data. While we do not present data on disease severity, our data corroborate with these findings by suggesting aberrant expression of monocyte subtypes in neurodegenerative disease. In addition, monocyte subtype redistribution may explain why total monocytes have not been found to be different in the MCI stage as compared to controls (42). Similarly, our identification of greater CD16+ NK cell overexpression in disease adds to existing literature identifying NK cell alterations in AD versus controls, however, this study did not compare CD16+ and CD16− NK cell subtypes (43). Peripheral myeloid DC have previously been found to be altered in AD and Parkinson’s disease versus controls (38,39). While results from PC2 suggest AD may indeed be characterized by a trend of reduced myeloid DC (Figure 3B), these cells did not strongly correlate to PC1 or PC3, thus underscoring the potential relative importance of monocytes and NK cells in all-cause neurodegenerative dementia. In all, our work builds upon extant immunophenotyping literature and confirms monocyte and NK cell population changes with multiple neurodegenerative diseases.

Monocytes are a heterogeneous population of innate immune cells. They have diverse functions, of cytokine release, migration to damaged tissues, and differentiation into phagocytic cells. Based on surface markers, monocytes can be divided into classical, nonclassical, and intermediate subtypes. Classical monocytes are pro-inflammatory, potentially neurotoxic, and are recruited to damaged tissue during inflammation (56). Nonclassical monocytes are anti-inflammatory and may be neuroprotective (57,58). In addition, monocytes may express functional receptors such as CCR2, a chemotaxis receptor controlling the recruitment of the cell, or TREM2, a receptor controlling phagocytic and inflammatory function (59,60). In contrast, NK cells in other diseases facilitate the death of infected cells, regulate the adaptive immune response through cytokine production, and mediate autoimmunity. NK cells can be subdivided based on their relative presentation of surface markers into CD16−, regulating cytokine production, or CD16+, indicating cytotoxicity (61). NK cells have been implicated in many disorders of the central nervous system, including neurodegenerative dementia (43,62,63).

Although previous evidence suggested monocyte, NK, and DC are each involved in neurodegenerative diseases, prior evidence was mainly gleaned from investigation in Alzheimer’s disease. Our results identify abnormal features of the peripheral innate immune system that are common across AD, LBD, FTD, CBS and PSP. By using data-driven analytic methods, we explore multivariate peripheral features that may not be revealed by evaluating one cellular type at a time. Innate immune patterns across the 19 cells investigated in this study identify patient groups to be dissimilar from controls (Figure 1). Applying a data-driven approach, our results imply neurodegenerative disease to be characterized by pro-inflammatory cytotoxic, peripheral immune dynamics atypical to healthy aging.

We observerd reduced PC1 loading in all-cause dementia patients versus controls (Figure 2A). This suggests that reduced relative expression of TREM2+ on peripheral monocytes and increased relative prevalence of classical monocytes may be a characteristic immune signature linked to neurodegenerative dementia. TREM2 is involved in modulating inflammation, mediating phagocytosis, and promoting myeloid cell survival (59,64). In the context of neurodegenerative diseases, TREM2 has recently been linked to prevention or downregulation of tau hyperphosphorylation (65). However, this interaction has been investigated only in microglia of murine models of AD rather than peripheral monocytes and human studies. Our results support claims of TREM2 as an important receptor acting from the periphery in neurodegenerative disease (27). Although the current study is correlational, without evidence of causality, the clinical relevance of the changes in innate immune profiles is highlighted by the associations of PC1 individual loading with survival across patient groups (Figure 4). Increased peripheral TREM2+ expression and nonclassical monocytes combined with reductions in classical monocytes and CD16+ NK cells may represent a protective effect in disease, while the opposite patterns may contribute to accelerated clinical decline. Moreover, the present link between increased TREM2+ monocytes and prognosis is especially relevant in light ongoing clinical trials targeting TREM2 in people with AD (66). Give the transdiagnostic loading of the immune profiles, TREM2 mediated treatments may be effective in other neurodegenerative conditions. In all, results warrant further investigation of TREM2+ monocytes, classical monocytes, and CD16+ NK cells.

Patients with LBD displayed lower median individual loadings in PC3 versus controls (Figure 4B). This third component was weighted positively by CD16− NK cells and negatively by CD16+ NK cells. Given CD16+ NK cells contributed to the negative arm of both PC1 and PC3, this cell type is of particular interest. Indeed, human post mortem studies show redistribution of NK cellular subtypes in patients with alpha-synuclein pathology, suggesting a potentially strong effect in this disease group (37,67). Reduced average PC3 loading in LBD suggests this cohort may harbor TREM2 dynamics dissimilar to other dementias as these cells comprise the negative end of PC3 (opposite to PC1 composition). The present findings distinguishing LBD are in line with previous observed differences in microglial activation in LBD as compared to other diseases (18,29).

Our study has several limitations. First, patient recruitment and cohort definition are based on clinical criteria rather than pathology-confirmed cases, although clinic-pathological correlations are high for PSP, FTD, LBD and AD. Next, the heterogeneity within cohorts (e.g., grouping bvFTD with PPA) and co-pathologies (e.g., CBS underpinned by CBD and/or AD) may complicate the interpretation of the results and reduce sensitivity to between-group differences. In addition, group sizes are unbalanced, although the non-parametric tests used are relatively robust to moderate variation in group size. Importantly, ACE-R was used as the primary assessment to capture cognitive decline as this exam is widely implemented in memory clinics. However, this screening-test varies in sensitivity to the domain-specific cognitive changes associated with different diagnoses, and this may have reduced power to detect correlations with cognitive deficits. Finally, data collection spanned several years, which may have led to batch variation in our immunophenotyping analysis. To help control sample variability, sample gating was conducted by a single person, and validated against the gating of two other experts on a sub-sample.

In conclusion, the present study provides a comprehensive characterization of the peripheral innate immune system in multiple neurodegenerative dementias. We suggest dysfunctional patterns of the innate immune system are characteristic of neurodegenerative diseases. Blood-derived innate immune profiles can distinguish sub-populations of cells relevant to diverse clinical cohorts and their prognosis. Further studies are needed to clarify interactions between the peripheral innate immunity profiles and dementia-related in the cerebrum. We hope that the identification of blood-based innate immune profiles can contribute to enhanced precision medicine approaches dementia, to identify new therapeutic targets and improve clinical trial design for immunotherapies.

## Data Availability

All data produced in the present study are available upon reasonable request to the authors

## Acknowledgments

This study was co-funded by the Dementias Platform UK and Medical Research Council (MC_UU_00030/14; MR/T033371/1); the Wellcome trust (103838; 220258); Race Against Dementia Alzheimer’s Research UK (ARUK-RADF2021A-010); the Cambridge University Centre for Parkinson-Plus (RG95450); the National Institute for Health Research (NIHR) Cambridge Biomedical Research Centre (BRC-1215-20014; NIHR203312: the views expressed are those of the authors and not necessarily those of the NIHR or the Department of Health and Social Care); the Progressive Supranuclear Palsy Association (PSPA2022/SMALL GRANTS002); the Addenbrookes Charitable Trust (Ref: 900380). For the purpose of open access, the authors have applied a Creative Commons Attribution (CC BY) license to any Author Accepted Manuscript version arising from this submission. This work is licensed under a Creative Commons Attribution 4.0 International License. We thank our participant volunteers for their participation in this study, thank the National Institute for Health Research (NIHR) Cambridge BioResource centre and staff, and the research nurses for their contribution, and the East Anglia Dementias and Neurodegenerative Diseases Research Network (DeNDRoN) for help with subject recruitment.

