## Supplementary Materials for "Peripheral innate immunophenotype in neurodegenerative disease: blood-based profiles and links to survival"

### Supplementary Information

### Cells and Flow Cytometry

#### Figure S1. Cell Ratios and Populations

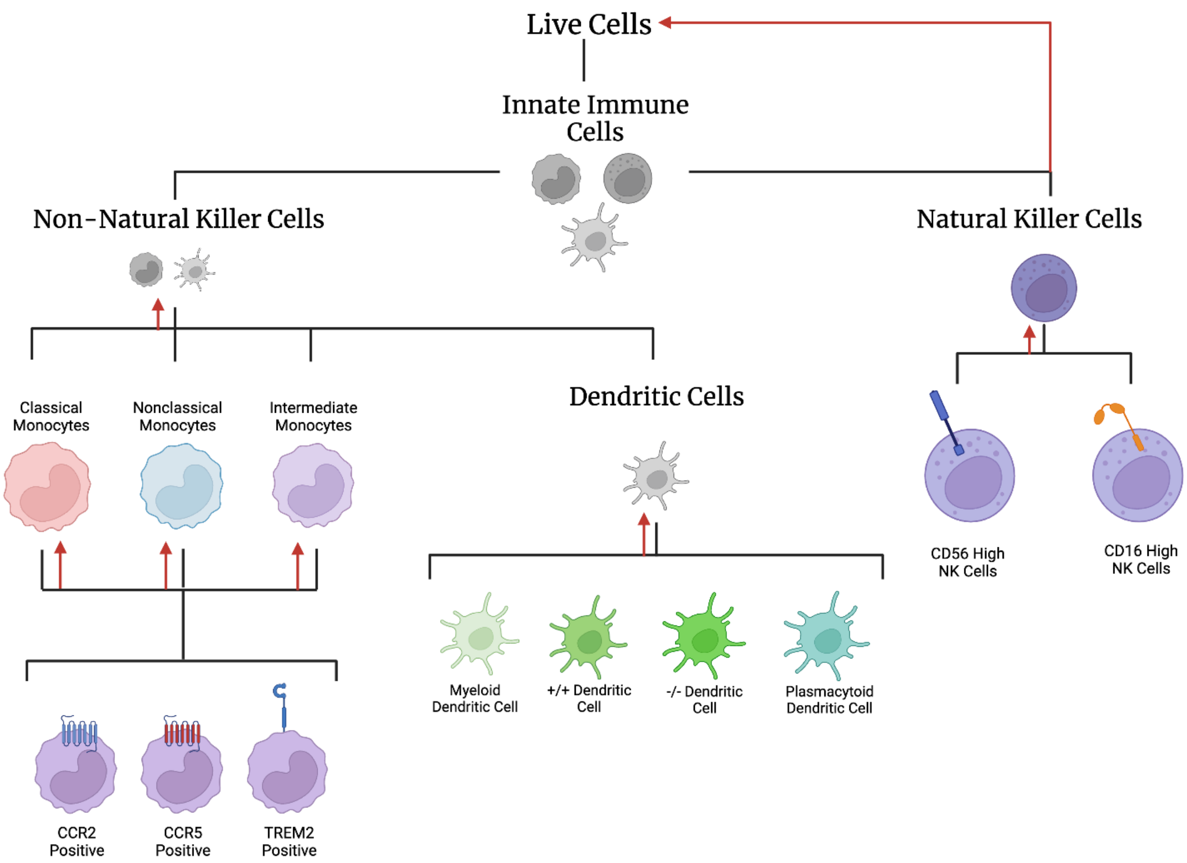

Figure 1. Graphic showing cell types in the study. Red arrows indicate the parent population that each raw cell below was divided by to yield the ratio value used for analysis. Grey populations were not considered in this analysis.

#### Figure S2. Gating Strategy

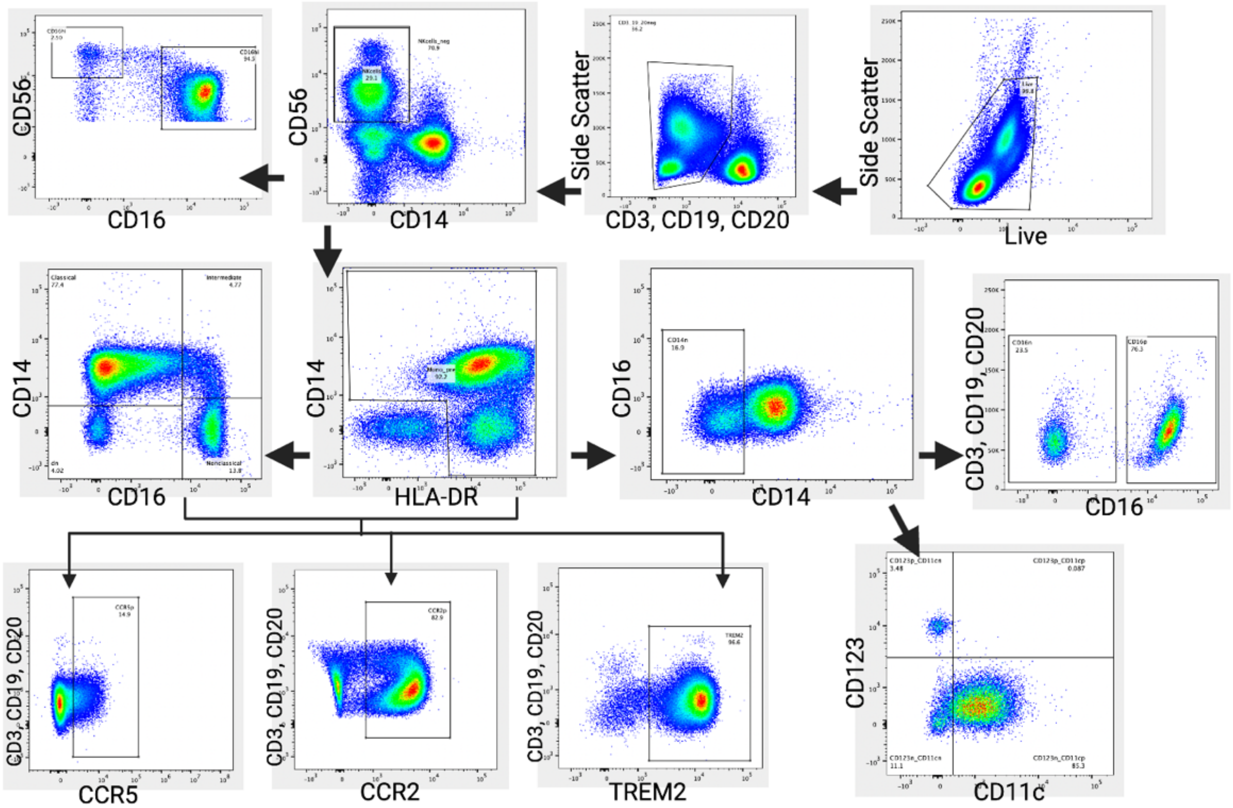

Figure S2. Gating Strategy for Flow Cytometry analysis. Samples were first evaluated for compliance by gating a continuous column flow over time. Next, all live cells were sectioned. B and T cells were eliminated by selecting cells negative for CD3, CD19, and CD20

#### Table S2. Antibodies, blocking reagent, and viability marker used in this study.

| Antibody | Vendor | Catalogue |
|---|---|---|
| CD56-FITC | eBioscience | 11-0566-42 |
| CD123-PerCP/Cy5.5 | eBioscience | 45-1239-42 |
| TREM2-PE<br>or<br>Rat IgG2B-PE (iso) | R&D Systems | FAB17291P<br>or<br>IC013P |
| CD11c-PE/Vio770 | Miltenyi Biotec | 130-113-581 |
| CCR5-AlexaFluor647 | eBioscience | 313712 |
| CD20-APC/Cy7 | eBioscience | 47-0209-42 |
| CD19-APC/Cy7 | eBioscience | 47-0199-42 |
| CD3-APC/Cy7 | BioLegend | 317342 |
| CCR2-BV421 | BioLegend | 357210 |
| CD14-BV605 | BioLegend | 301834 |
| CD16-BV650 | BioLegend | 302042 |
| HLADR-BV785 | BioLegend | 307642 |
| Live/Dead Aqua | Thermo Fisher | L34957 |

**Table S2: Antibodies, blocking reagent, and viability marker used in this study**

#### ICC Results

##### Table S3.

| Gate | ICC Value |
|---|---|
| Live | 0.96 |
| Innate Cells | 0.96 |
| NK Cells | 0.95 |
| Non-NK Cells | 0.97 |
| Dendritic Cells | 0.95 |
| Classical Monocytes | 0.54 -> 0.91 |
| Intermediate Monocytes | 0.9 |
| Nonclassical Monocytes | 0.12 -> 0.9 |

Table S3. ICC Results across major gates. The gating strategy was validated between three distinct raters using the same gating strategy individually for 10 shared samples. Inter-rater correlation was assessed graphically as well as mathematically using intraclass correlation coefficient (ICC) analysis: ICC 2,1 (Shrout and Fleiss, 1979). ICC values for parent populations are found in Table 1. Graphical representations of inter-rater correlation can be found in the appendix (see figures Figure 1.1 – 1.25). Classical Monocyte and Nonclassical Monocyte inconsistencies were evaluated and normalized prior to continuing with the analysis. The ICC value upon adjustment is also reported.

Figure S3. Visual Verification of Gating Strategy.

10 patients were selected and gated by three distinct raters. In addition to ICC analysis, gates that were visually inconsistent (see below) were revisited and the strategy was agreed upon between the three raters before continuing with analysis. (E.g. Gater 3 had consistently different values past figure 1.23 which were addressed).

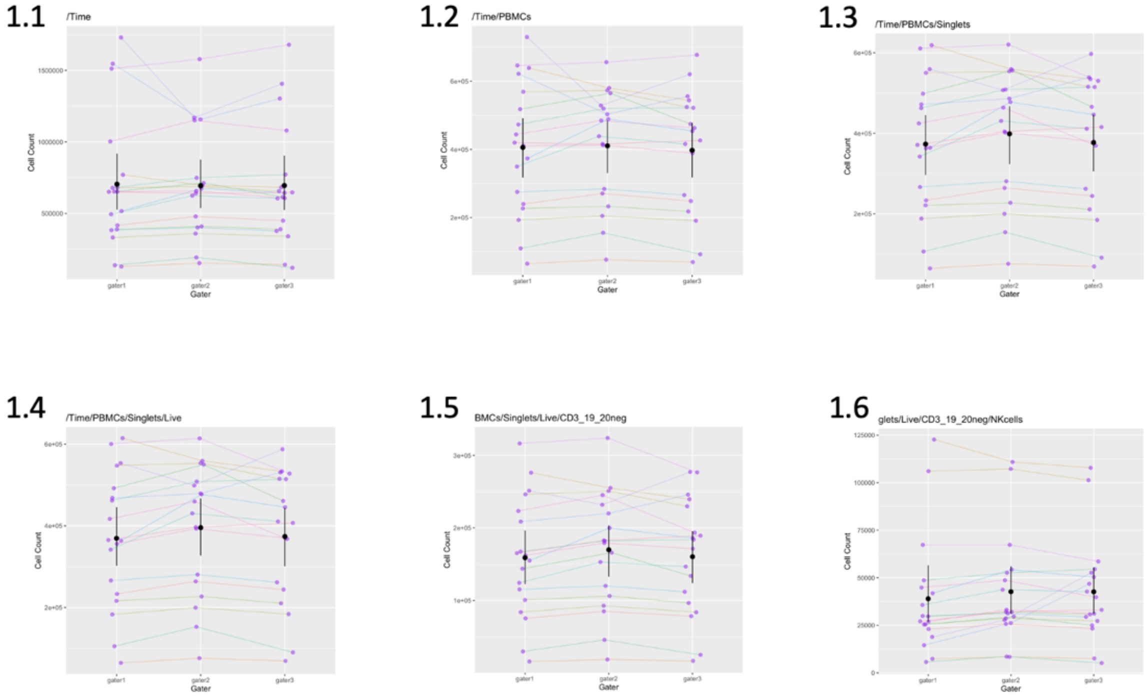

1.7

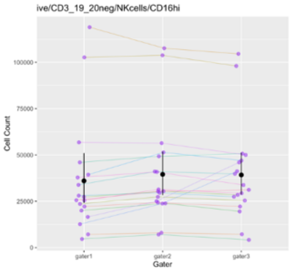

1.8

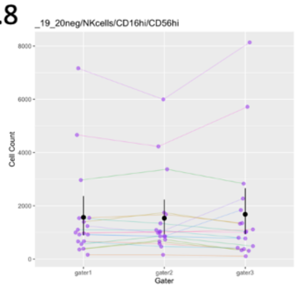

1.9

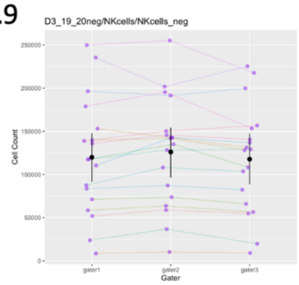

1.10

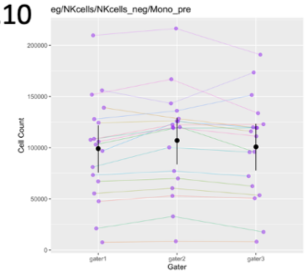

1.11

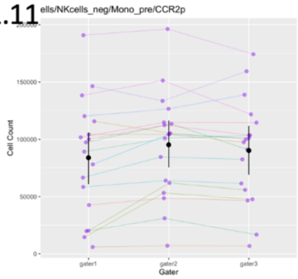

1.12

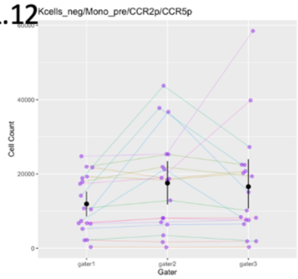

1.13

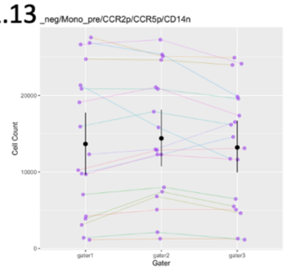

1.14

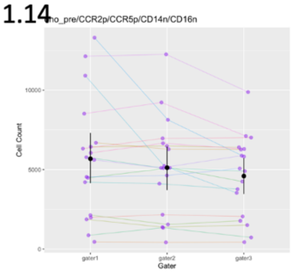

1.15

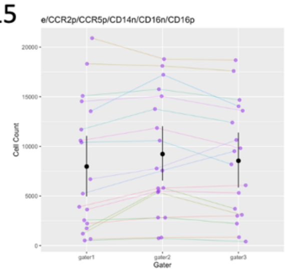

1.16

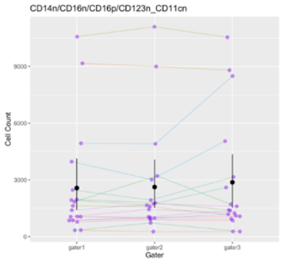

1.17

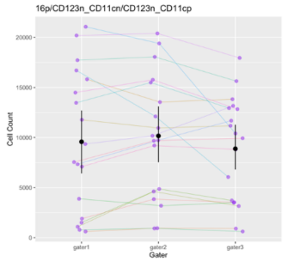

1.18

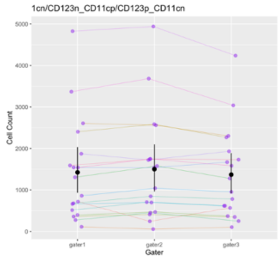

1.19

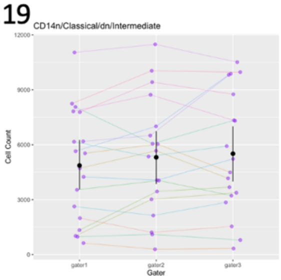

1.20

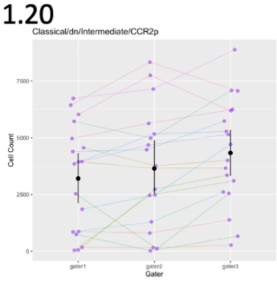

1.21

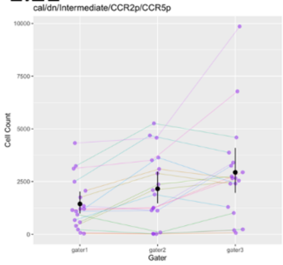

1.22

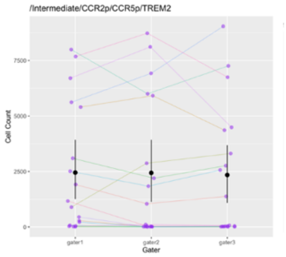

1.23

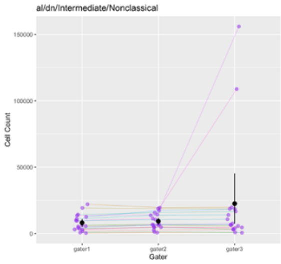

1.24

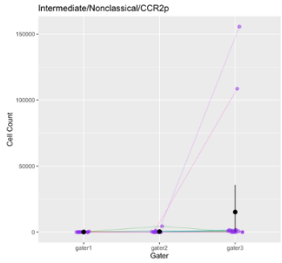

1.25

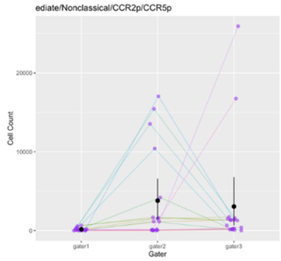

1.26

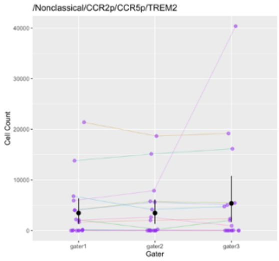

1.27

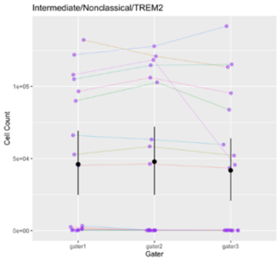

### PCA Results

#### Table S4. PC3 Post Hoc analysis Results

| | | p | p_fdr |
|---|---|---|---|
| Control | AD | 0.04* | 0.426 |
| Control | CBS | 0.44 | 1 |
| Control | LBD | 0.212 | 0.01* |
| Control | FTD | 0.0163 | 0.212 |
| Control | PSP | 0.31 | 1 |
| AD | CBS | 0.245 | 1 |
| AD | LBD | 0.286 | 1 |
| AD | FTD | 0.634 | 1 |
| AD | PSP | 0.185 | 1 |
| CBS | LBD | 0.0228 | 0.273 |
| CBS | FTD | 0.121 | 1 |
| CBS | PSP | 0.957 | 1 |
| LBD | FTD | 0.63 | 1 |
| LBD | PSP | 0.00668 | 0.094 |
| FTD | PSP | 0.0816 | 0.816 |

Table S4. Results for Dunn's Post hoc analysis in individual PC3 loading. Results for comparisons between each group individual loading in PC3in the three components selected. * Indicates significance at $p < 0.05$.

#### Table S5. Cell Percent Contribution to each selected component

Each cell type percent contribution to principal component 1, 2, and 3.

| Cell Type | PC1 | PC2 | PC3 |
|---|---|---|---|
| TREM2+ Classical Monocytes | 17.78 | 0.99 | 5.06 |
| TREM2+ Nonclassical Monocytes | 17.43 | 2.31 | 7.21 |
| TREM2+ Intermediate Monocytes | 15.36 | 3.04 | 8.66 |
| Classical Monocytes | 11.25 | 1.57 | 3.09 |
| Nonclassical Monocytes | 9.14 | 0.38 | 3.52 |
| CCR2+ Nonclassical Monocytes | 7.93 | 1.25 | 0.88 |
| CD16+ NK Cells | 4.93 | 0.21 | 20.08 |
| CCR2+ Classical Monocytes | 4.49 | 11.24 | 0.09 |
| DC +/+ | 3.41 | 0.41 | 0.72 |
| DC -/- | 2.79 | 13.70 | 1.14 |
| CCR5+ Classical Monocytes | 2.04 | 3.71 | 2.72 |
| Intermediate Monocytes | 1.07 | 0.24 | 11.14 |
| CD56+ NK Cells | 0.82 | 0.00 | 16.35 |
| CCR2+ Intermediate Monocytes | 0.66 | 15.26 | 0.75 |
| NK Cells | 0.35 | 0.04 | 15.23 |
| CCR5p Intermediate Monocytes | 0.27 | 17.40 | 0.25 |
| CCR5+ Nonclassical Monocytes | 0.14 | 15.57 | 0.65 |
| Plasmacytoid DC | 0.10 | 0.27 | 2.43 |
| Myeloid DC | 0.03 | 12.41 | 0.02 |

#### Figure S4. Scree Plot

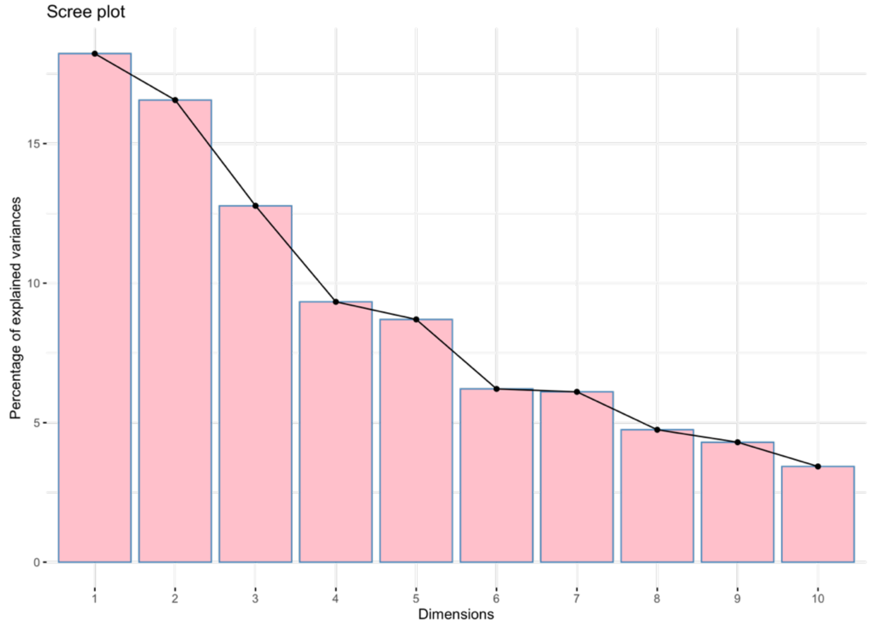

Figure S3. Scree plot from PCA computed following the removal of outliers.

#### Figure S5. LOF Histogram Visualization

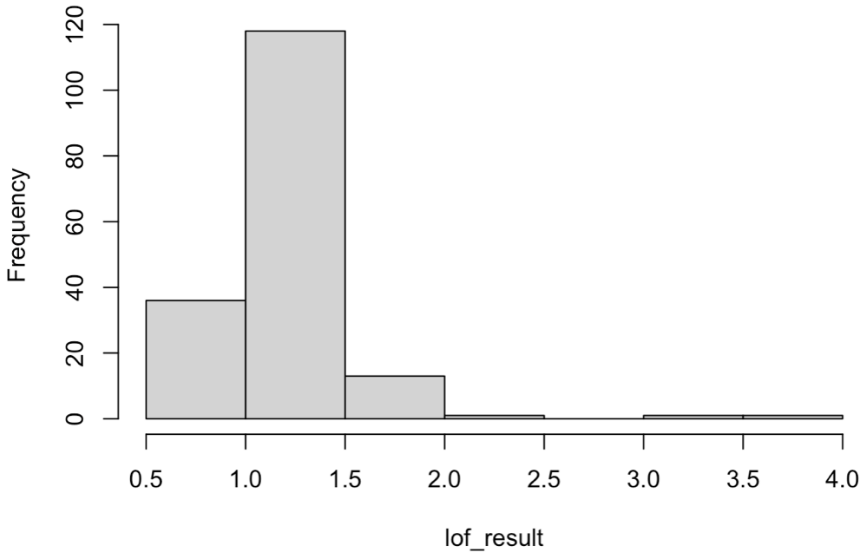

Figure S4. Histogram visualization of LOF- 2 subjects with LOF > 3 were removed prior to PCA recomputation.

Table S5. PC Comparisons to Age, Sex, and ACE- R

| | PC1 | PC2 | PC3 |
|---|---|---|---|
| Age Interaction Statistics<br><br>Statistic, p-value | f= 0.026,<br><br>p = 0.70 | f= 0.1237,<br><br>p= 0.725 | f= 0.3254,<br><br>p= 0.5691 |
| Sex Interaction Statistics<br><br>Statistic, p-value | U=3739,<br><br>p = 0.281 | U= 4243,<br><br>p = 0.742 | U= 3598,<br><br>p = 0.141 |
| ACE-R Correlation Statistics<br><br>statistic, p-value | t= 0.6024,<br><br>p =0.439 | t =0.436,<br><br>p = 0.663 | t= -0.780,<br><br>p= 0.436 |

#### Individual Cells: Pairwise Comparisons

##### Table S6. Results from Analyses of Dementia and Controls

| Cell Type | p_value | p_fdr | statistic |
|---|---|---|---|
| CCR2+ Classical Monocytes | 0.0939 | 1 | 2265 |
| CCR5+ Classical Monocytes | 0.463 | 1 | 2541 |
| TREM2+ Classical Monocytes | 0.759 | 1 | 2847 |
| CCR2+ Intermediate Monocytes | 0.943 | 1 | 2778 |
| CCR5+ Intermediate Monocytes | 0.97 | 1 | 2745 |
| TREM2+ Intermediate Monocytes | 0.493 | 1 | 2958 |
| CCR2+ Nonclassical Monocytes | 1.74E-04** | 0.003306* | 3857.5 |
| CCR5+ Nonclassical Monocytes | 0.961 | 1 | 2741.5 |
| TREM2+ Nonclassical Monocytes | 0.376 | 1 | 3016 |
| DC-/- | 0.743 | 1 | 2853 |
| Myeloid DC | 0.878 | 1 | 2711 |
| Plasmacytoid DC | 0.216 | 1 | 3120 |
| DC+/+ | 0.835 | 1 | 2695 |
| CD16+ NK Cells | 0.051 | 0.969 | 2184 |
| CD16- NK Cells | 0.0203* | 0.3857 | 3437 |
| Classical Monocytes | 0.00118** | 0.02242* | 1805 |
| Intermediate Monocytes | 0.278 | 1 | 3075 |
| Nonclassical Monocytes | 0.00134** | 0.02546* | 3697 |
| NK Cells | 0.00879** | 0.16701 | 1988 |

Table S6. Results for pairwise comparisons of each cell type between dementia patients and controls. * Indicate significance at $p < 0.05$. See Figure 6 for graphical representation of values.

#### Figure S6. Individual Cell types between controls and dementia

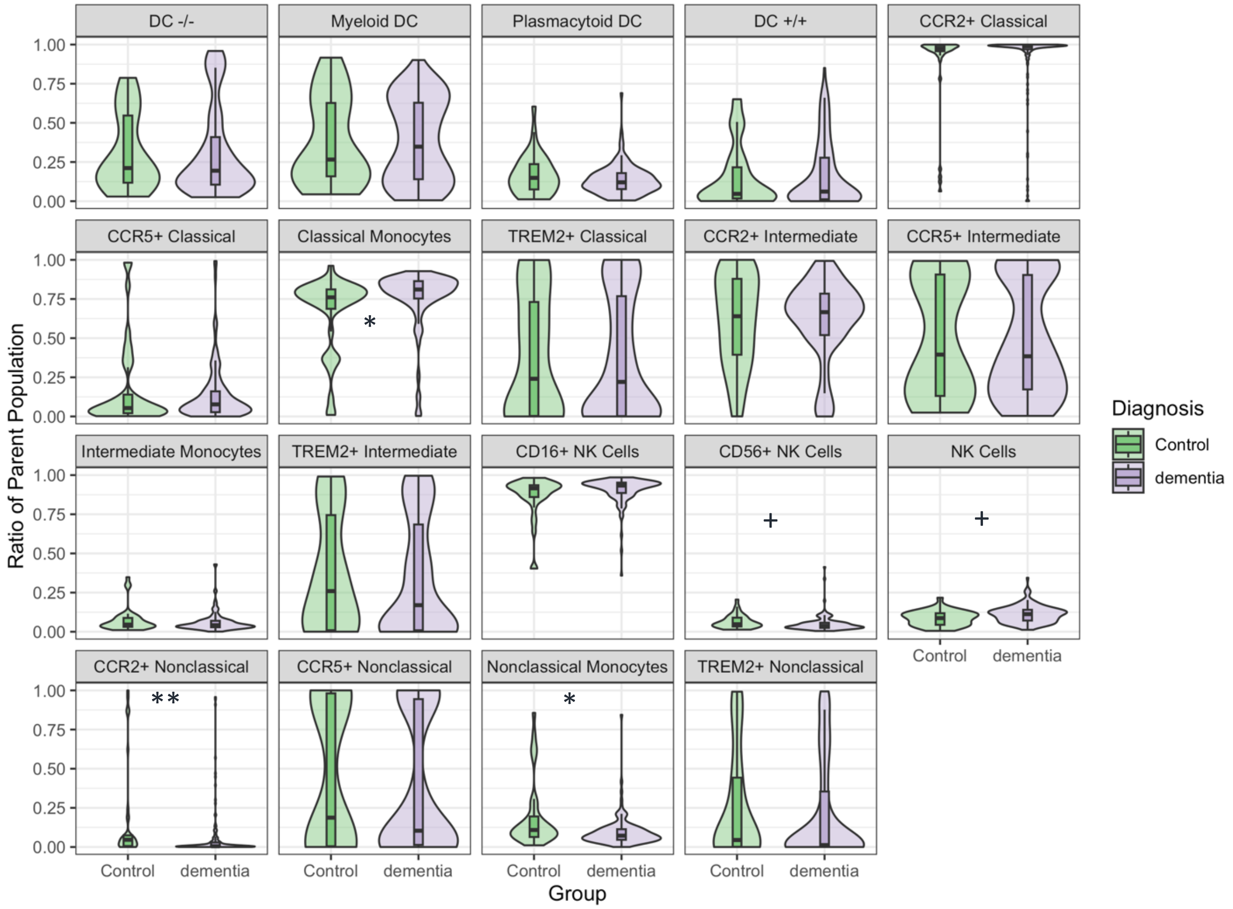

Figure S6. Median prevalence of innate immune cells (as a ratio of parent) between dementia patients and controls. * indicates statistical significance following FDR correction, + indicated statistical significance prior to FDR correction.
